# Cross-Cultural Adaptation of Scales Measuring Stigma Related to HPV, HIV, and Cervical Cancer Stigma for Use in a Kenyan Context

**DOI:** 10.1101/2025.08.02.25332873

**Authors:** Amber C. Smith, John A. Gallis, Yujung Choi, Elizabeth A. Bukusi, Breandan B. Makhulo, Melissa H. Watt, Joao R.N. Vissoci, Megan J. Huchko

## Abstract

**Objective:** To develop and validate a culturally adapted stigma scale to measure the levels and effects of human papillomavirus (HPV)- and cervical cancer-related stigma, especially as cervical cancer prevention strategies become globally available.

**Methods:** Between February 2020 and May 2022 in Kisumu, Kenya, we adapted items from existing validated scales addressing HIV, HPV, and cervical cancer stigma and incorporated additional items based on qualitative data, forming an item pool across three dimensions: stigmatizing attitudes, enacted stigma, and internalized stigma. Cognitive interviews (CIs) were conducted in English and Dhluo to refine items. Psychometric properties were assessed using exploratory and confirmatory factor analysis.

**Results:** Following CIs, we conducted a pilot survey with 998 participants and subsequently a validation survey with 480 women. Psychometric analysis identified 77 items with satisfactory properties across four health domains. The average factor loading of retained items was 0.64 (SD = 0.08), and all final scale models demonstrated high internal consistency (Cronbach’s alpha > 0.9).

**Conclusions:** This rigorously validated tool offers a reliable method to evaluate stigma’s impact on screening behaviors and assess stigma-responsive interventions as HPV vaccination and testing expand globally.

**Clinical Impact Statement:** Our team developed contextually relevant item pools to measure stigma related to HPV and cervical cancer in areas of high HIV prevalence in western Kenya. Through cognitive interviews, pilot survey, and validation survey, we tailored down the final item pool to 77 items with Cronbach’s alpha all greater than 0.9. As HPV vaccination and testing become more available in the target location, it remains imperative for researchers and providers to recognize and avoid furthering stigma. These tools can be used to measure the impact of stigma on health engagement and outcomes and the impact of stigma-reduction interventions in this area.

## INTRODUCTION

While Human Papillomavirus (HPV) vaccination and testing have the potential to nearly eliminate cervical cancer in high-income settings in the next century, low-and middle-income countries (LMICs) have not had equal access to or uptake of these technologies (Geneva: World Health Organization, 2020). In 2020, 604,127 women developed and 341,831 died of cervical cancer (The Global Cancer Observatory, 2021), with almost nine out of ten cases and deaths occurring in LMICs (Abbas et al., 2020; Fitzmaurice & Global Burden of Disease Cancer Collaboration, 2018; Soerjomataram et al., 2012). In Kenya, where cervical cancer is the most common cancer among women, the crude incidence rate is 19.4 cases per 100,000 women (Bruni et al., n.d.; Ngutu & Nyamongo, 2015), over 2.5 times the rate in the United States (*American Cancer Society | Cancer Facts & Statistics*, n.d.). Availability and accessibility of the highly effective HPV-based prevention technologies are essential to combat the burden of cervical cancer. Both vaccination and screening for HPV have recently been recommended by the Kenya Ministry of Health in their cancer control strategy. In planning for implementation of these programs, work must be done to understand the personal beliefs and social factors that influence the uptake of HPV-related services.

Knowledge and beliefs play a significant role in personal risk perception and health behaviors. One potential barrier to the uptake of screening or vaccination is stigma related to either cervical cancer or HPV, which may be experienced differently in areas affected by high rates of HIV and HIV-related stigma. Stigma and discrimination correlate with both poor physical and mental health outcomes and can manifest in different forms, including fear-based stigma, value-based stigma, enacted stigma, and internalized stigma (Feyissa et al., 2019; Hatzenbuehler et al., 2013). Researchers have shown a clear association between HIV-related stigma and health behaviors and outcomes, including uptake of HIV testing, adherence to medication and psychological well-being (Endeshaw et al., 2014; B. Turan et al., 2017; Vanable et al., 2006).

HIV-associated stigma is well-characterized, with a recent systematic review identifying at least 12 distinct validated instruments across different languages to measure HIV-related stigma among people living with HIV (PLWH) (Relf et al., 2021). Cervical cancer, like HIV, can be highly stigmatized, impacting treatment follow-up and engagement (Endeshaw et al., 2014; Feyissa et al., 2019; Ginjupalli et al., 2022; Vanable et al., 2006). There has been little work rigorously measuring HPV-related stigma and its possible impact on attitudes toward HPV-based cervical cancer prevention (Geneva: World Health Organization, 2020). HIV and HPV are both sexually transmitted viruses, and women living with HIV (WLWH) have an increased risk for HPV, all potential “markers” of stigma. Lessons from HIV-related stigma research highlight the importance of using validated tools to measure stigma, which will be crucial to the development and testing of interventions seeking to impact HPV-related stigma. This is of paramount importance as HPV-based prevention is central to the WHO’s strategy for elimination of cervical cancer as a global health problem and its “90-70-90” targets to meet by 2030.

Understanding the magnitude and impact of stigma related to HPV, HIV, and cervical cancer requires a framework describing the factors associated with stigma and its impact on health behaviors, as well as a reliable, validated tool to measure stigma levels. While other researchers have adapted HIV stigma scales for East Africa, to our knowledge, this has not been done for HPV or cervical cancer (Sao et al., 2022; Visser et al., 2008). To develop an effective tool for the Kenyan context, our team created the “HPV-Related Stigma Framework” to describe the relationship between the potential stigmas related to these three health domains, identified possible impacts on health beliefs and behaviors, and proposed modifiers of both the stigma and the behavior impact (Ginjupalli et al., 2022). The framework incorporated findings from qualitative work with women and community health volunteers in western Kenya which suggested that misinformation and lack of understanding of the relationship between HPV and cervical cancer were some of the key drivers of HPV-related stigma at the individual level. It also incorporated societal drivers, including religious and cultural beliefs. These factors together were felt to decrease personal risk perception and negatively affect prevention behaviors (Ginjupalli et al., 2022).

While HIV-related stigma scales have been validated in multiple settings, the available scales for HPV and cervical cancer have been developed and deployed in high-income countries. To our knowledge, there are no item pools that examine the interrelationship of the three health domains to describe intersectional stigma (Holzemer et al., 2007; Marlow & Wardle, 2014; Neufeld et al., 2012; J. M. Turan et al., 2011; Visser et al., 2008; Waller et al., 2007). In Western Kenya, a culturally adapted item pool would improve the understanding of HPV and cervical cancer-related stigma among WLWH and in areas of high HIV prevalence. We used the HPV-Related Stigma Framework to develop an item pool in English and Dhluo, a language local to western Kenya, to measure stigma related to the three health domains of cervical cancer, HIV, and HPV. This paper describes the process of item development and psychometric testing to create a validated measurement tool for stigma, which can be used to assess the impact of stigma on health behaviors evaluate stigma-focused interventions to promote uptake of HPV-based cervical cancer prevention.

## METHODS

### Overview

We conducted a three-phase study to develop and psychometrically test an item pool to measure HPV, cervical cancer, and HIV-Related Stigma (Fig 1). In Phase I, we developed the item-pool and carried out cognitive interviews to determine content validity. In Phase II, we evaluated internal structure and reliability, identifying items to be maintained into the confirmatory phase. In Phase III, we confirmed the internal structure through surveys collected on an independent sample.

**Figure 1:**
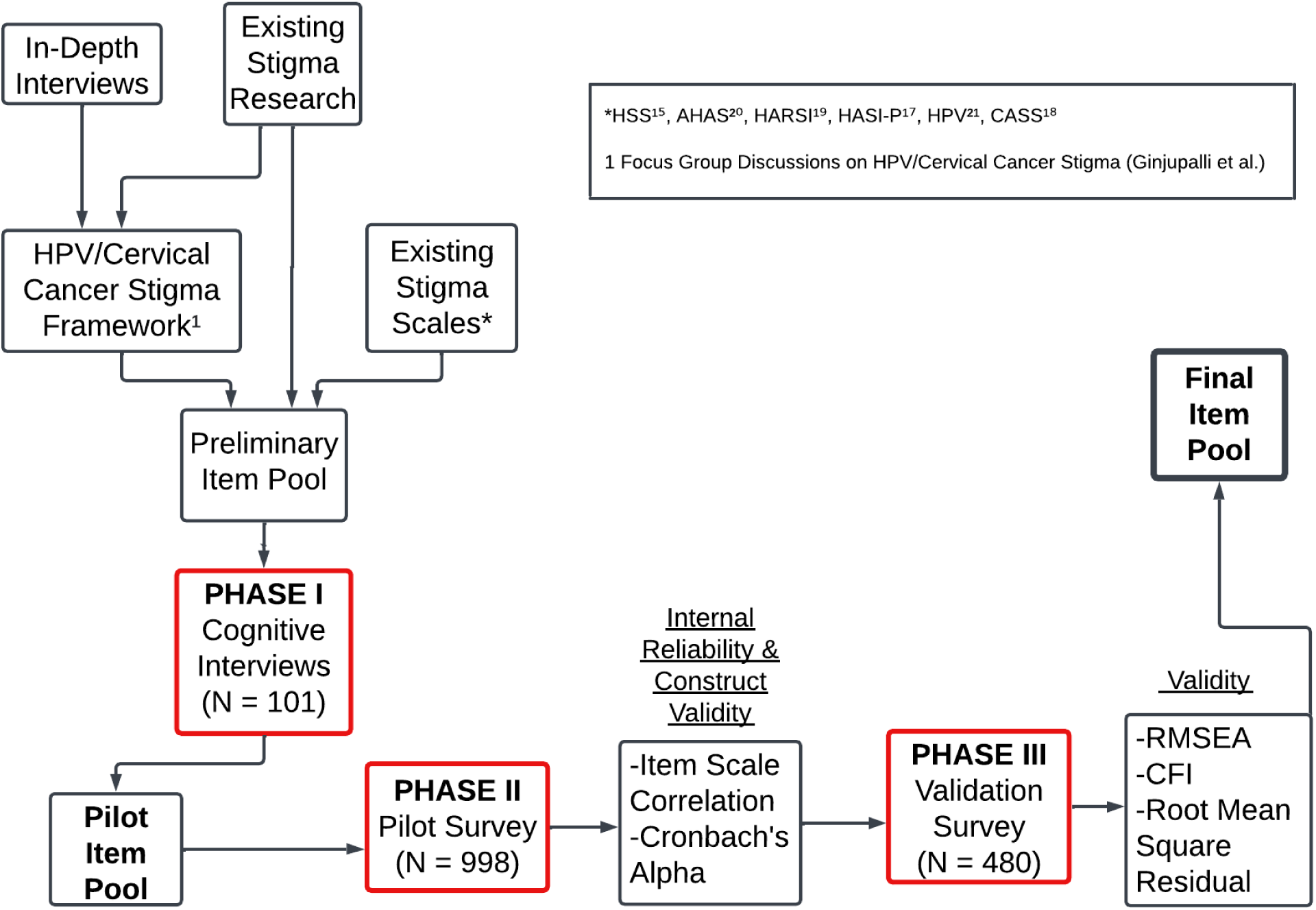
Item Development and Evaluation of Psychometric Properties for Measurement Tool for HPV, Cervical Cancer, and Hiv-Related Stigma.

### Setting

We recruited participants for all phases from Lumumba Sub-County Hospital, a high-volume, government-supported health center in Kisumu, Kenya providing a variety of inpatient and outpatient services. Women living in Kisumu who were 18 years and older and able to provide informed consent were approached and screened in the waiting area and then led to a private area for informed consent, enrollment, and data collection.

### Phase I. Evaluating evidence of validity related to test content

#### Item Pool Development

To develop the item pools, we used the HPV-Related Stigma Framework, along with qualitative data from formative work in this population (Ginjupalli et al., 2022) and existing literature (Fig 1) to adapt items from existing scales (Holzemer et al., 2007; Marlow & Wardle, 2014; Neufeld et al., 2012; J. M. Turan et al., 2011; Visser et al., 2008; Waller et al., 2007) or create items to cover context-specific aspects of the scale (Table 1). Items were brief statements that could be answered on a 4-point Likert scale (strongly disagree, disagree, agree, or strongly agree). To understand the multiple types of stigma, we sought a balanced number of items to measure blame and judgement, interpersonal distancing, shame, and anxiety for each of the health domains. Because of the relatively recent introduction of HPV testing in the region, many women would not have had access to testing and therefore we phrased items in the future tense to capture anticipated stigma, either internalized or enacted.

**Table 1:** Items Adapted and Created About HPV, HIV, and Cervical Cancer Across the Stigma Dimensions.

| Health Domain |  | Stigma Dimensions |  |  |  |  |  |
| --- | --- | --- | --- | --- | --- | --- | --- |
|  |  | Attitudes | Anticipated Internalized^ | Anticipated Enacted | Total Items | Examples of Some Items Created and Adapted: |  |
|  |  |  |  |  |  | Original Item from Existing Scale | Item Adaptation |
| HPV |  | 15 (2) | 17 | 19 | 51 | I am ashamed that I'm HIV positive.(Neufeld et al., 2012) | I would be ashamed to have HPV. |
|  |  |  |  |  |  | N/A | Women with HPV should not get pregnant. |
| Cervical Cancer |  | 11 | 12 (1) | 11 | 34 | If people knew my HIV status, I would be treated badly by health workers.(J. M. Turan et al., 2011) | If I had cervical cancer, I would be treated badly by health workers. |
|  |  |  |  |  |  | N/A | If I had cervical cancer, I would be ashamed by my symptoms (bleeding, weight loss) |
| HIV | HIV Negative | 20 (2) | 16 | 12 | 48 | If people knew my HIV status, my family would reject or abandon me. (J. M. Turan et al., 2011) | If I had HIV, my family would reject or abandon me. |
|  |  |  |  |  |  | N/A | Women with HIV should not be mothers. |
|  | WLWH | * | 16 ** | 12 | 28 | I feel completely worthless.(Holzemer et al., 2007) | Having HIV makes me feel completely worthless. |

(N) = number of new items developed from focus group discussion data
WLWH = Women Living with HIV
\* = To reduce potential additional stigma for WLWH, we did not ask questions about stigmatizing attitudes
<sup>^</sup> = Anticipated Internalized categorizes items where an individual who may not necessarily know their status or is negative anticipates how they would feel towards their self if they had that disease. *For example, "If I had HPV, I would feel guilty for getting infected with HPV" (HPV13).*
\*\* = Internalized

Between February and October 2020, we conducted cognitive interviews (CIs) with 101 women. The CI guide included seven questions covering understanding, comfort, and cultural fit of items in English and Dhluo (Supplementary 4). Participants were also asked for any suggestions to improve wording or clarity. We sought a minimum of at least five participants to evaluate each item. Items were split across the health domains of HPV, cervical cancer, and HIV (separated for respondents who were living with HIV compared to HIV Negative). For the HIV-related items, we altered wording for participants who identified as WLWH and women who identified to be HIV-negative and did not include the items about stigmatizing attitudes to avoid additional trauma or stigma for these women already living with the disease. As the HIV-related items had already been validated in English among people from East Africa these items were translated and only assessed among Dhluo-speaking women, serving as an external validation set. Participants had the choice to conduct interviews in either English or Dhluo based on their preference, and the interview was limited to approximately 10 to 15 items from a single domain to reduce fatigue. There was a total of 51 HPV-related items and 34 cervical cancer-related items. There were 48 survey items for HIV-negative women and 28 items for WLWH across anticipated enacted and internalized. Results from the CIs were used to edit the item pools for further testing for reliability and validity.

### Phase II. Evaluating the evidence of reliability, dimensionality and item’s fit to the measures

Next, exploratory factor analysis (EFA) techniques were used to determine reliability, dimensionality, and item-fit between January and February 2022. For the exploratory factor analysis, we aimed for each item to be assessed by 7 participants, as has been done in prior studies (Terwee et al., 2007). We determined that at least 980 participants would need to participate to assess items in English and Dhluo.

To achieve precision around the factor levels while minimizing distortions from interfactor correlations, model area, secondary loadings and unequal loadings, we used the formula of seven participants per item to determine the sample size (Wolf et al., 2013). The principal-factor method was used to analyze the correlation matrix. We first examined eigenvalues and percent of variation explained by factors to determine the number of factors to retain for each scale. Items with relatively large factor loadings (≥0.5) and no large cross-loadings (other factor loadings <0.3 if examined factor loading is between 0.5 and 0.6, or other factor loadings <0.4 if examined factor loading is ≥ 0.6) were tentatively included in the final scale for the next stage.

After exploratory factor analysis, the partial credits method from item response theory was used to further refine the items. We examined a person-item map to determine overlap between levels of items and removed items with a relatively low amount of space between levels. Next, we examined the outfit and infit statistics, and removed items with outfit or infit > 1.2 (Wu et al., 2016). Finally, for simplification purposes the items selected on each scale were made to be the same between English and Dhluo versions of the scale.

### Phase III. Evaluating evidence of validity related to internal structure and to other variables

We conducted validation testing of the full item pool using data from an additional 480 women between April and May 2022 (Mundfrom et al., 2005). Confirmatory factor analysis (CFA) was used to verify the construct validity of the instrument through: a) item-factor parameter and item-items individual reliability, b) absolute, incremental and parsimonious fit indexes; and c) average variance extracted to examine the convergent validity (De Vellis, 2003; Dishion et al., 2007). The fit of the CFA models was evaluated using the comparative fit index (CFI), root mean square error of approximation (RMSEA) 90% CI upper bound, and the standardized root mean square residual (SRMR) at their standard cutoffs (Hu & Bentler, 1999). If the fit indices fell out of the standard range, we further examined modification indices.

We then explored external validity through correlation testing between answers to final item pool and the validated HIV and AIDS Stigma Instrument for People Living with HIV (HASI-P) (Holzemer et al., 2007). Additionally, we used Cronbach’s alpha to assess internal reliability of the scales (Terwee et al., 2007). Again, the same items were removed or retained in both English and Dhluo for scale consistency.

For the confirmatory factor analysis, we used guidelines from Wolf et al. (2013) (Terwee et al., 2007; Wolf et al., 2013). Given the three scales (English, Dhluo for WLHW, and Dhluo for HIV-) with 2-3 factors each (HPV, CC, and HIV+/-), with about 6-8 or more items loading on each factor, and conservatively assuming low factor loadings, we would need 160 participants per scale to be adequately powered. Hence, the needed sample size for the validation survey was 480. We also have 90% power to detect a Pearson correlation of 0.25 or higher between the newly created stigma scale and another validated stigma scale.

#### Ethical statement

The study protocol was reviewed and approved by both the Duke Health Institutional Review Board (Reference ID: 30362) and the Kenya Medical Research Institute Scientific and Ethics Review Unit (Protocol No. 3894). All participants provided written informed consent.

## RESULTS

### Phase I

The initial item pool consisted of 161 questions (Table 1). Almost all items, 156, were derived from existing scales and 5 were newly created based on data from Focus Group Discussions. 101 women completed CIs, with each participant assessing an average of 13.2 items, with each item assessed by an average of 5.4 participants. The average participant age was 30.3 years (range 19 to 51). Of the interviews, 64 were conducted in Dhluo and 37 were conducted in English. 68 women had previously been screened for cervical cancer and 31 women were self-reported to be living with HIV, which was evenly represented across interviews in Dhluo (19/64, 29.7%) and English (12/37, 32.4%) (Supplementary 1).

There were 22 items evaluated further after at least two participants said the item should be eliminated (Supplementary 2). The reasons for suggested elimination were mainly related to the potential stigma of the question and included fear or judgement (9 questions), causing pain or making women feel uncomfortable or embarrassed (6 questions), difficulty understanding (1 question), and other reasons of perceived unacceptability (6 questions). For example, one respondent recommended eliminating *People with HIV have only themselves to blame for getting HIV* (Visser et al., 2008) “because it would bring them pain and self-blame,” utilizing descriptors that allude to stigma (Supplementary 2). Almost all suggestions for elimination came from the Dhluo participants, with 20 of the suggested 22 items, including 11 from HPV, 4 from HIV, and 5 from cervical cancer. Among the CIs in English, 2 of the HPV items were flagged. No questions among the WLWH cohort (only tested in Dhluo) were suggested for follow-up and none among the cervical cancer items tested in English were flagged. Overall, one of 5 (20%) newly developed items and 21 of 156 (13.5%) adapted items were flagged for further investigation.

After the CIs, most items were acceptable for further testing, either as presented or with revised translations. Only two items were eliminated: “I would feel irritated by someone with cervical cancer” and “I would feel anxious about getting cervical cancer” were both eliminated due to team concerns about redundancy to other items and participant fatigue with the survey length. The team also decided to adapt three acceptable items from the cervical cancer screening to both HPV and HIV, adapt an HIV item to HPV health domain, and add a previously validated item for WLWH. Therefore, the item pool going into EFA had 168 items, with 54 HPV items, 32 cervical cancer items, 50 items for HIV negative women, and 32 items for WLWH (Supplementary 3).

### Phase II

Following the CIs, 998 women in Kisumu, Kenya completed the pilot survey consisting of a subset of items for reliability and construct validity testing (Table 2). The average participant age was 31.3 (range 18 to 65). Of the women’s surveys, 606 (60.7%) were conducted in Dhluo, and 392 (39.3%) were conducted in English. 471 women (47.4%) had previously been screened for cervical cancer and 401 (42.1%) were self-reported to be living with HIV (Supplementary 1).

**Table 2:**
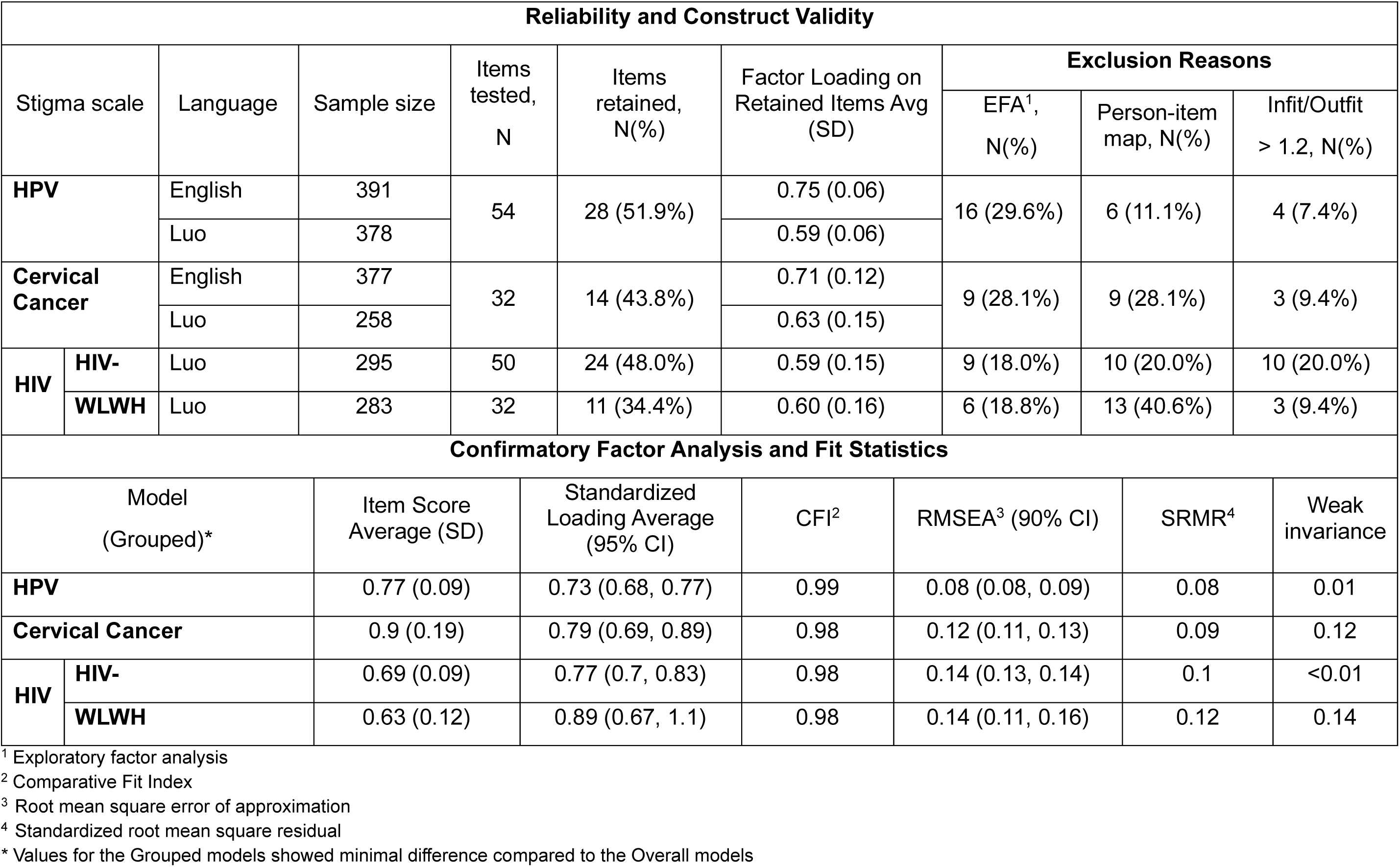
Pilot Test of Preliminary Item Pool to Measure HPV, Cervical Cancer, and HIV-Related Stigma Among 998 Women, Confirmatory Factor Analysis and Scale Characteristics, & Fit Statistics.

From the item pool, 28 of 54 (51.9%) HPV items were retained, 14 of 32 (43.8%) on the cervical cancer stigma scale, 11 of 32 for WLWH, and 24 of 50 for HIV-negative women after the pilot testing phase. A total of 91 (54.2%) of the 168 items in the pilot item pool were removed prior to confirmatory factor analysis (Supplementary 3). The majority excluded items at this stage were removed based on the factor loading in the EFA being less than 0.3 or examining the Graded Partial Credit Model’s person-item map. The average factor loading among remaining items was 0.64 (0.08). Cronbach’s alpha was estimated to be >0.9 for all scale models in phase II.

### Phase III

For Phase III, 480 women completed the validation survey consisting of the edited pool of 77 items. The average participant age was 34.0 (range 20 to 70). Of the surveys, 300 (62.5%) were conducted in Dhluo, with the rest (180, 37.5%) conducted in English. 317 women (66.0%) had previously been screened for cervical cancer and 137 (28.5%) were self-reported to be living with HIV (Supplementary 1).

Confirmatory Factor Analysis indicated factor loadings for each domain in a very good range, with the average standardized factor loading greater than 0.7. Table 2 displays the fit statistics for the overall models (invariant to language) and multiple groups model (grouped by language). The relative fit as measured by the CFI is above 0.95 in all cases. The RMSEA upper 90% CI and SRMR indicate adequate fit for all models. Thus, all items were retained as part of the final scales. Modification indices were examined, but freeing up item covariances did not substantively improve the fit or the conclusion.

Cronbach’s alpha for all final scale models were all greater than 0.9. The Pearson and Spearman correlations of the newly created scales with the HASI-P were all relative strong (>0.60), indicating good evidence of validity regarding relationship with other variables (Table 3). The final item pool consisted of 28 items related to HPV, 14 related to cervical cancer, and 24 translated items related to HIV for external validity testing. The HIV items including 13 probing discriminatory attitudes which were not administered to WLWH (Table 4). Supplementary 3 displays the flow of the number of items from cognitive interview phase to final item pool.

**Table 3:** Overall Mean Scores of HPV, Cervical Cancer, and HIV Stigma Scales and Correlation with HASI-P.

| Health Domain* |  | English<br>(n = 180) | Dhluo<br>(n = 300) | Total<br>(n = 480) | HASI-P Correlation |  |
| --- | --- | --- | --- | --- | --- | --- |
|  |  |  |  |  | Pearson | Spearman |
| **HASI-P\*\*** |  | 0.34 (0.47) | 0.66 (0.56) | 0.54 (0.55) |  |  |
| **HPV** |  | 0.68 (0.52) | 0.82 (0.54) | 0.77 (0.54) | 0.598 | 0.605 |
| **Cervical Cancer** |  | 0.81 (0.53) | 0.96 (0.59) | 0.90 (0.58) | 0.604 | 0.614 |
| **HIV** | **HIV Negative Women** | 0.51 (0.48) | 0.81 (0.56) | 0.70 (0.55) | 0.779 | 0.757 |
|  | **Women Living with HIV** | 0.63 (0.52) | 0.65 (0.54) | 0.64 (0.53) | 0.734 | 0.761 |

\*All items are coded on a 0, 1, 2, and 3 scale, with a lower score indicating less endorsement of stigma.
\*\* HIV and AIDS Stigma Instrument for People Living with HIV

**Table 4:**
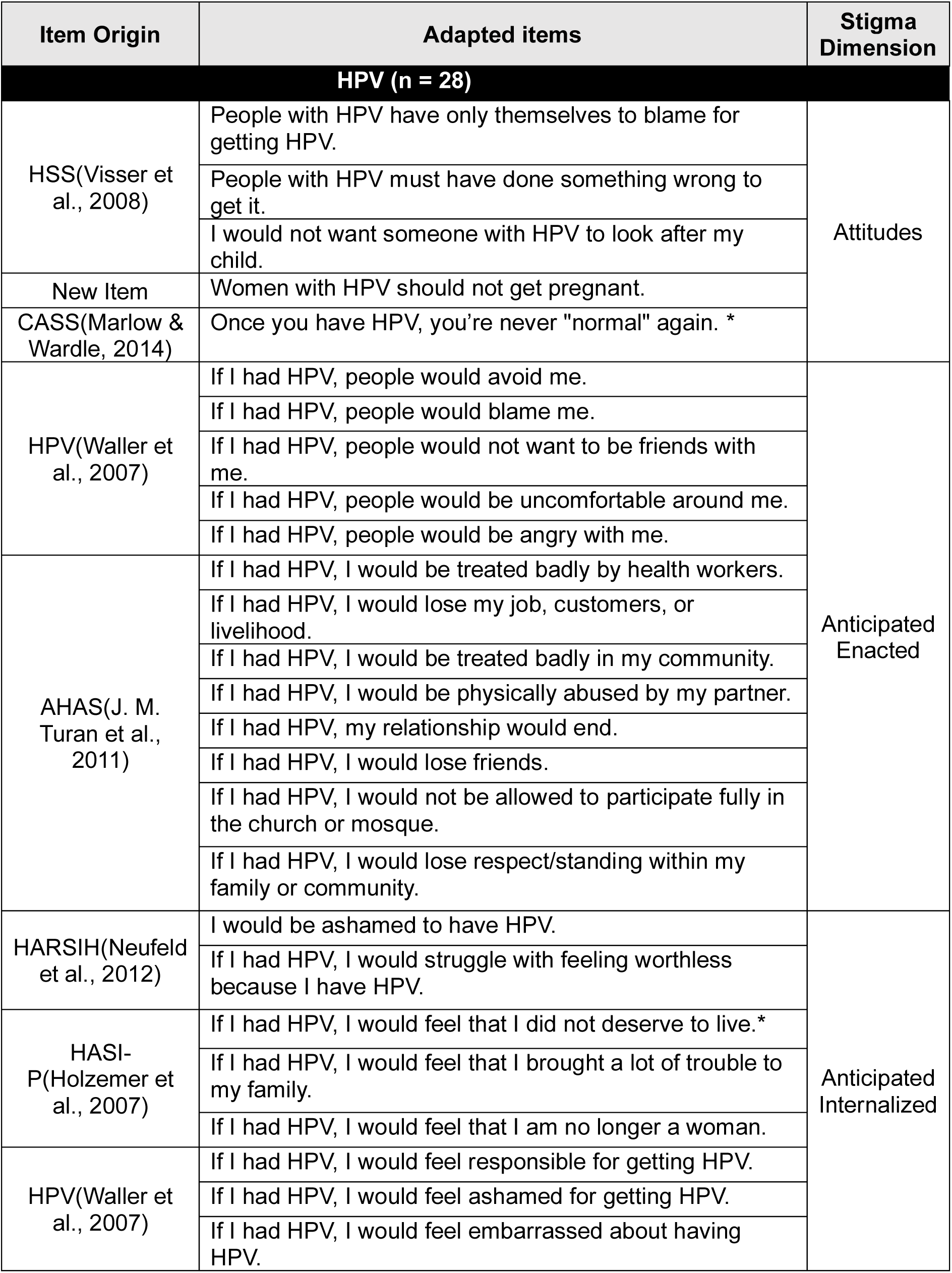
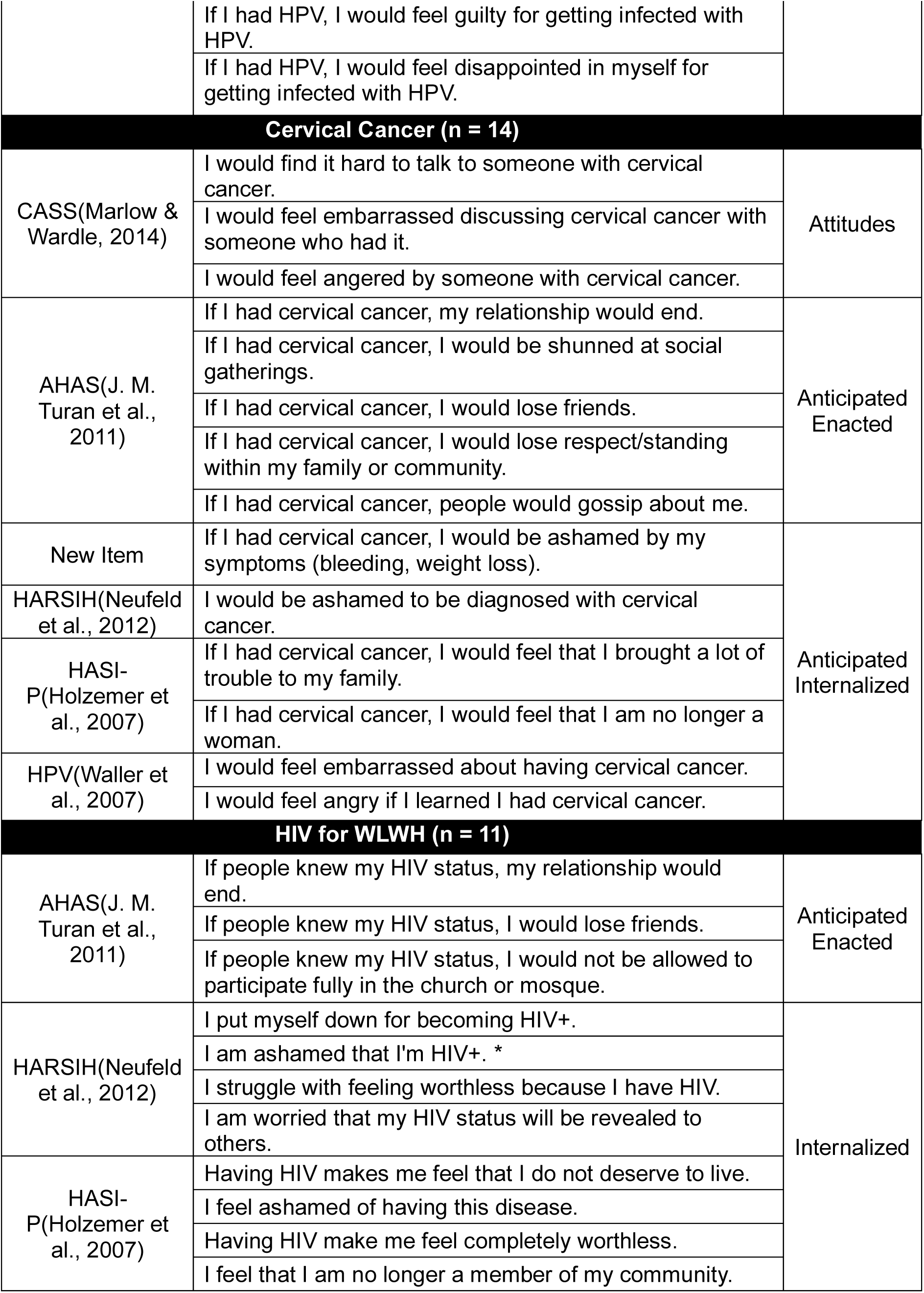
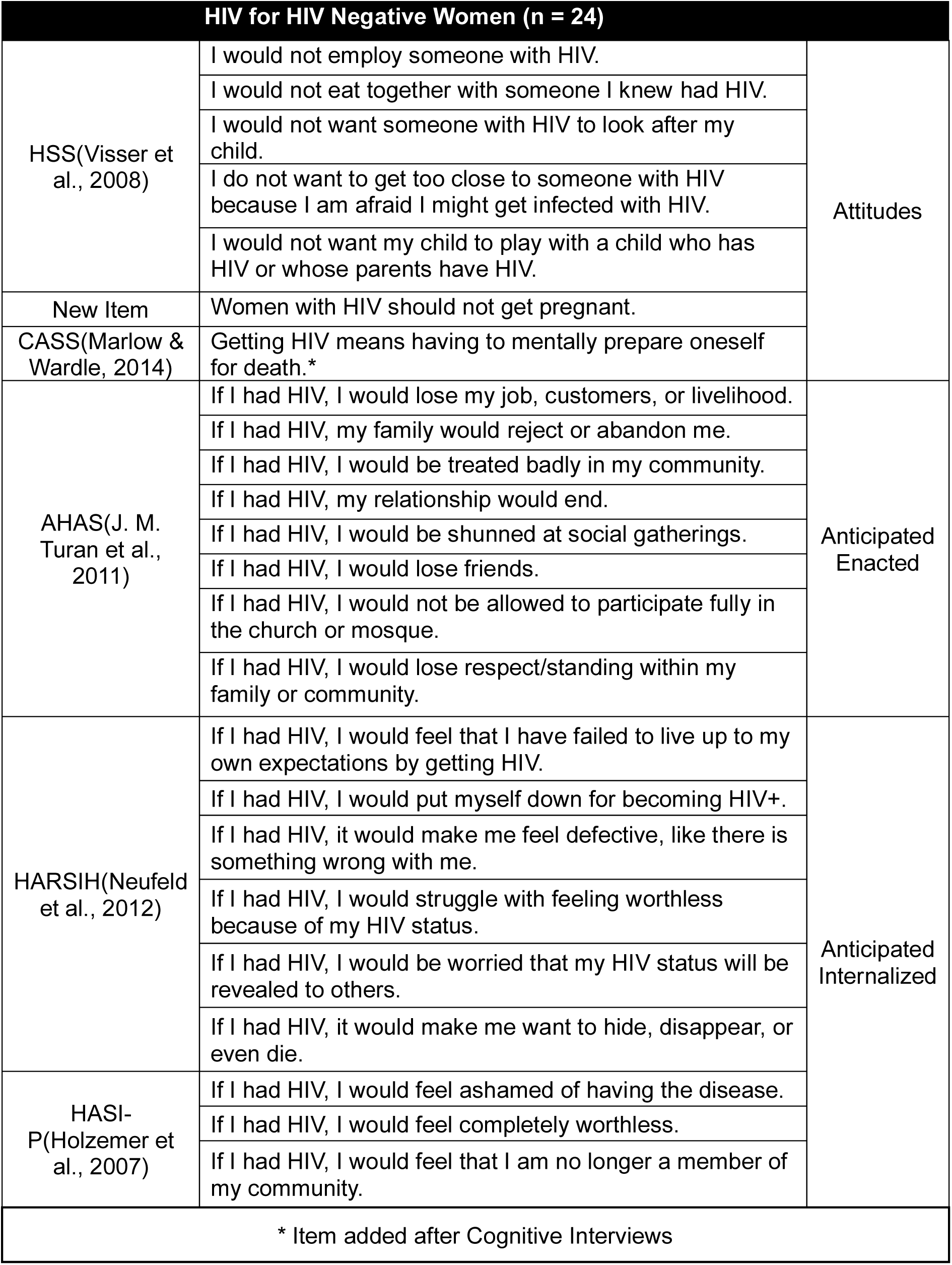
Final Item Pool for HPV, Cervical Cancer, and HIV-Related Stigma After Dropping Items after Cognitive Interview, Pilot Survey, and Validation Survey.

## DISCUSSION

We used data from qualitative work and the HPV-related Stigma Framework (Ginjupalli et al., 2022) to identify and adapt items from existing scales (Holzemer et al., 2007; Marlow & Wardle, 2014; Neufeld et al., 2012; J. M. Turan et al., 2011; Visser et al., 2008; Waller et al., 2007) and derive additional items specific to HPV and cervical cancer. Cognitive interviews among English and Dhluo speaking women found that most of the items were clear and acceptable, and further testing for reliability and validity helped to reduce the overall number of items for the final item pool. We sought to develop an item pool to measure levels of stigma related to HPV and cervical cancer for women in Western Kenya to help guide programmatic and educational strategies for HPV-based cervical cancer prevention.

While almost all the items tested through cognitive interviews were acceptable, the most common reasons that participants suggested elimination of items were their potential to cause or worsen stigma. For example, cognitive interviewers suggested elimination utilizing words like “pain and self-blame,” “brings fear,” and “kill my ego”, suggesting the potential presence of HPV stigma already within Kenya (Gabbidon & Chenneville, 2021; Maestre et al., 2018). Almost all suggestions for elimination came from Dhluo participants, indicating issues with translation or factors in their educational and cultural backgrounds that impacted their experience with or perception of stigma. Understanding key distinctions among different populations who may be vulnerable to these stigmas is essential when considering how to introduce and integrate screening and educational programming. This underscores the importance of exploring both the translation issues and the distinct educational and cultural factors that may be unique to women’s experiences to ensure that the survey is accurate, culturally appropriate, and does not inadvertently introduce stigma in this population (Choi et al., 2012; Villagran, 2022).

This quantitative measure of stigma related to HPV, cervical cancer, and HIV among women was found to have satisfactory psychometric properties. As is common in measure developments, the initial item pool was reduced by a large extent. The final group of items retained showed adequate evidence of validity related to internal structure, other variables and evidence of reliability. While we initially sought a distribution of items representing across stigma domains (exacted, anticipated, internalized) and constructs (shame and blame, social distancing) these were not maintained during the validation process. This reflects the tension between the utility of these concepts to define and understand the layers of stigma, and the complexity of the lived experience. While the item pool was developed to be comprehensive, it likely falls short of capturing the multi-dimensionality of internalized, anticipated, or experienced stigma.

In addition to inherent limitations in quantifying a complex and multi-dimensional personal experience, there are other limitations we must note. Our study population may not reflect the general population in the region, with the vast majority of participants in the CIs having completed secondary education and over half some kind of post-secondary education, three to five times the rates in the general population (Supplementary 1) (Kenya National Bureau of Statistics, 2019). As education correlates with stigma, we may have under-sampled women more vulnerable to stigma, which may more accurately reflect the population.

This item pool can provide an objective and context-specific means to measure the relationship between stigma and screening behaviors, as well as the impact of stigma-responsive interventions. Future work could identify and measure intersectional stigma to assess the overlap and interplay between drivers of stigma within HIV, HPV and cervical cancer, and the distinct lived experiences for women with these conditions. Next steps include deploying the scale to assess levels and demographic correlates of stigma across these health domains to consider the necessity of item weighting, as well as to look at the association with cervical cancer prevention behaviors. As HPV-based prevention strategies begin to scale, this is a unique opportunity to utilize this rigorously developed item pool to understand pre-existing stigma and avoid stigmatizing messaging that could impact uptake of interventions.

## Data Availability

All data produced in the present study are available upon reasonable request to the authors.

