## Supplemental Figures for "Cross-Cultural Adaptation of Scales Measuring Stigma Related to HPV, HIV, and Cervical Cancer Stigma for Use in a Kenyan Context"

**Title**

### Supplementary Figures

| **Supplementary 1:** Sociodemographic Characteristics of Participants in the three testing groups: Cognitive Interview, Pilot Survey, and Validation Survey | | | |
|---|---|---|---|
| | Cognitive Interview, n = 101 n(%) | Pilot Survey, n = 998 n(%) | Validation Survey, n = 480 n(%) |
| ***Age*** | *n = 101* | *n = 994* | *n = 480* |
| *Age Range* | *19 - 51* | *18 - 65* | *20 - 70* |
| 18-25 Years | 30 (29.7%) | 236 (23.7%) | 43 (9.0%) |
| 26-34 Years | 45 (44.6%) | 465 (46.8%) | 250 (52.1%) |
| 35-70 Years | 26 (25.7%) | 293 (29.5%) | 187 (39.0%) |
| ***Education*** | *n = 101* | *n = 994* | *n = 480* |
| Primary or none | 11 (10.9%) | 183 (18.4%) | 221 (46.0%) |
| Secondary | 36 (35.6%) | 499 (50.2%) | 202 (42.1%) |
| Post-Secondary | 54 (53.5%) | 312 (31.4%) | 57 (11.9%) |
| ***Cognitive Interview Language*** | *n = 101* | *n = 998* | *n = 480* |
| English | 37 (36.6%) | 392 (39.3%) | 180 (37.5%) |
| Dhluo | 64 (63.4%) | 606 (60.7%) | 300 (62.5%) |
| **Living with HIV** | *n = 101* | *n = 952* | *n = 480* |
| WLWH | 31 (30.7%) | 401 (42.1%) | 137 (28.5%) |
| **Previously Screened for Cervical Cancer** | *n = 101* | *n = 994* | *n = 480* |
| Yes * | 68 (67.3%) | *471 (47.4%)* | 317 (66.0%) |

* Women responding "don't know" were included within the sample calculation denominator, but they were excluded from the category of being screened previously

**Supplementary 2:** Items Suggested by Cognitive Interviewers for Elimination Across Assigned Category of Reasons

| Assigned Category for Suggested Elimination | Item | Cognitive Interview Language | Quotes of Reasons for Suggested Elimination from Cognitive Interviewers * |
|---|---|---|---|
| **HPV Health Domain** | | | |
| Difficulty Understanding | If I had HPV, people would think I was unclean.[21] | English | "It doesn't make sense"; "Because it makes someone uncomfortable" |
| Painful, uncomfortable, or embarrassing | If I had HPV, people would not want to have a sexual relationship with me.[21] | English | "Because it is embarrassing"; "Because it will kill my ego if I have HPV and the survey is conducted on me" |
| Painful, uncomfortable, or embarrassing | Getting HPV is a punishment for bad behavior.[15] | Dhluo | "It makes women feel uncomfortable"; "It is kind of passing judgement on someone that they have bad behavior" |
| Painful, uncomfortable, or embarrassing | Women with HPV should not get pregnant * | Dhluo | "Because it is an uncomfortable question to answer"; "It is uncomfortable" |
| Fear or judgement | I feel uncomfortable around someone with HPV.[15] | Dhluo | "Because it might scare someone who doesn't know about HPV"; "Because maybe both the participant and interviewer could be pos and the response given would not sound good" |
| Fear or judgement | I would not want someone with HPV to look after my child.[15] | Dhluo | "It is so judgmental" |
| Fear or judgement | I do not want to get too close to someone with HPV because I am afraid I might get infected with HPV.[15] | Dhluo | "Because it might bring fear in them"; "It can make women have stigma" |
| Fear or judgement | If I had HPV, people would think badly of me.[21] | Dhluo | "It will instill fear and they might not screen"; "It is judgmental" |
| Fear or judgement | If I had HPV, people would blame me.[21] | Dhluo | "Because it will make them fear to be judged"; "It is judgmental" |
| Fear or judgement | If I had HPV, people would be angry with me.[21] | Dhluo | "Because it can discourage women from screening with fear of being disliked if they turn positive for HPV" |
| Other | If I had HPV, my family would reject or abandon me.[20] | Dhluo | "It gives a picture that HPV is a terrible thing"; "Because people might never tell the truth" |
| Other | If I had HPV, people would gossip about me.[20] | Dhluo | "Because it is a normal thing in our society"; "I feel if people don't know my status they would not gossip" |
| Other | If I had HPV, I would feel responsible for getting HPV.[21] | Dhluo | "It is very personal and sensitive"; "It is too harsh" |
| **Cervical Cancer Health Domain** | | | |

| | | | |
|---|---|---|---|
| Fear or judgement | Getting cervical cancer means having to mentally prepare oneself for death.[18] | Dhluo | "It will bring fear to women"; "It is traumatizing to talk about death" |
| | Having cervical cancer would make me want to hide, disappear, or even die.[19] | | "It brings fear"; "Words used are very strong" |
| | If I had cervical cancer, I would feel that I did not deserve to live.[17] | | "It can bring fear"; "It can bring down their self-esteem" |
| Painful, uncomfortable, or embarrassing | I would feel guilty for having cervical cancer.[21] | | "It is a disturbing question"; "Because nonwoman can ask to have cervical cancer" |
| | I would feel irritated by someone with cervical cancer.[18] ** | | "Because it will bring out the suffering that cervical cancer patients undergo" |
| **HIV Negative Health Domain** | | | |
| Painful, uncomfortable, or embarrassing | People with HIV have only themselves to blame for getting HIV.[15] | Dhluo | "Because it would bring them pain and self-blame"; "It is accusing them falsely for what is not their fault" |
| Other | Getting HIV is a punishment for bad behavior.[15] | | "It will make women have deep thoughts and remind them how they contracted HIV"; "It is not fair for those who were maybe born with it"; "It is too straight forward and would not bring out the answer we are looking for" |
| | I would be troubled if someone with HIV moved in next door to me.[15] | | "Because nothing can happen to you even if your neighbor is sick"; "Because people can get help" |
| | If I had HIV, people would gossip about me.[20] | | "Because that is normal, and people will always gossip"; "In case she heard someone talk about her status and then said about this she can be stressed or depressed it would remind her what she heard someone say about her" |

*Responses from interviewers that were repeated almost verbatim from another interviewer were omitted from the table.*

* Item newly developed following Cognitive Interviews

** Eliminated after Cognitive Interview

**Supplementary 3:** Item Pool Size Across All Health Domains Throughout Cognitive Interviews, Pilot Survey, and Validation Survey

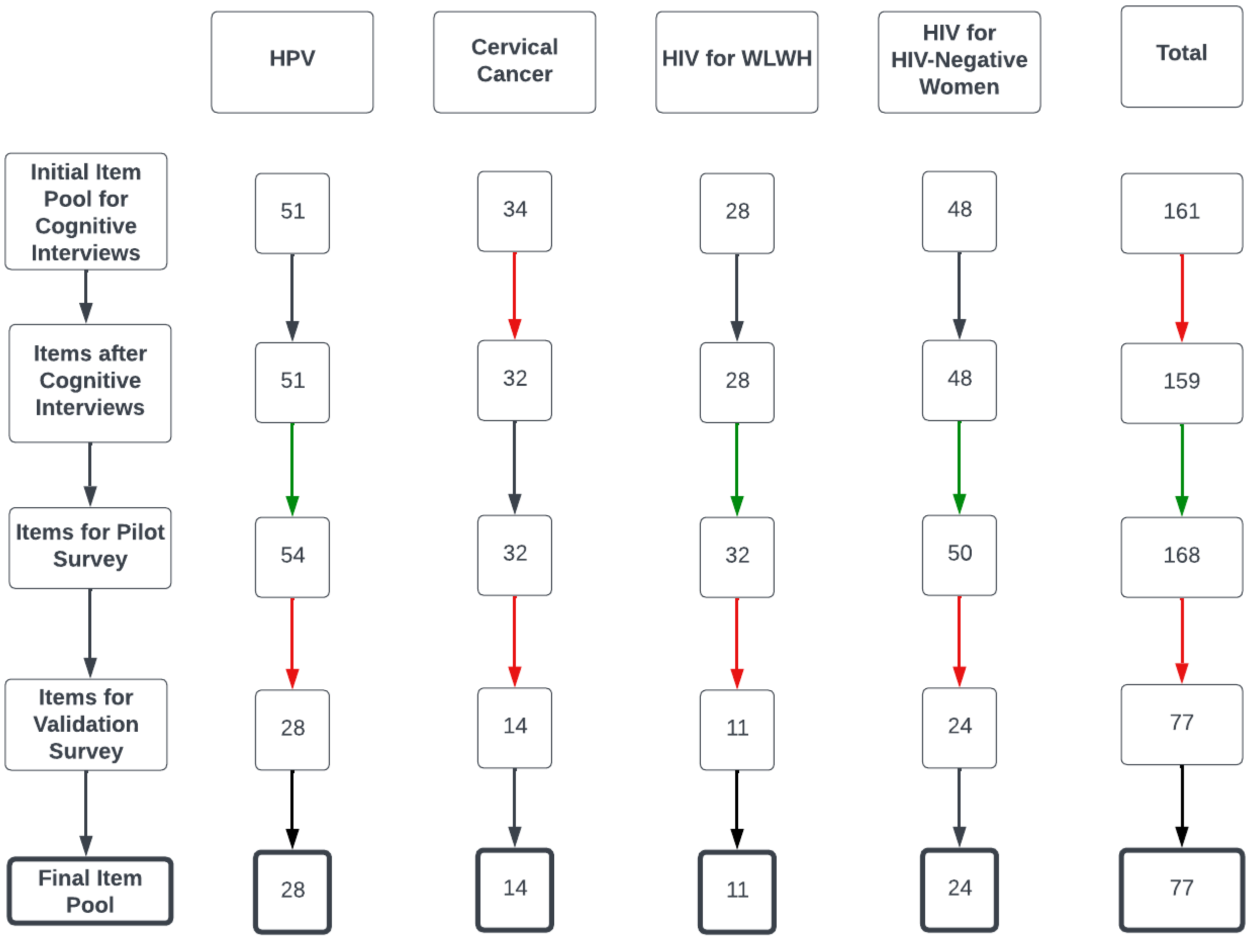

KEY:

*Green arrows indicate added items.*

*Red arrows indicate removed items.*

*Black arrows indicate no change in item pool number.*

**Supplementary 4:** Questions Asked of Cognitive Interviewers for each Assigned Item

| Question | Responses |
|---|---|
| (1) Did you understand this question? | * |
| (1b) Please, tell us in your own words what the question is asking. | Explain |
| (2) Did you have any difficulty using the response options? | * |
| (3) Were there any words or ideas you did not know/understand? | * |
| (4) Do you think the questions should be simplified? | * |
| (5) Would this question make women feel uncomfortable? | * |
| (5b) Please explain, regardless of Y/N response | Explain |
| (6) Would women feel like they could answer this question truthfully? | * |
| (6b) If yes, please explain | Explain |
| (7) Do you think this question should be eliminated? | * |
| (7b) If yes, why should it be eliminated? | Explain |
| (7c) If no, why not? | Explain |

* Answer options = Yes, No, Don't Know, Declined to Answer

| **Supplementary 5:** Summary of the HIV Stigmatizing Items with Sources and Modifications | | | | | | | |
|---|---|---|---|---|---|---|---|
| | | Women Living with HIV | | | HIV Negative Women | | |
| **Item Origin** | **Item Adapted** | **Item Modification-Women Living with HIV** | **Item Kept** | **Cognitive Interview** | **Item Modification-HIV-Negative Women** | **Item Kept** | **Cognitive Interview** |
| ***Attitudes*** | | | | | | | |
| HSS[15] | Getting HIV is a punishment for bad behavior. | | | | Getting HIV is a punishment for bad behavior. | | ** |
| | I would think less of someone if I found out the person has HIV. | | | | I would think less of someone if I found out the person has HIV. | | |
| | I would be upset if someone with HIV moved in next door to me. | | | | I would be troubled if someone with HIV moved in next door to me. | | ** |
| | I feel uncomfortable around someone with HIV. | | | | I feel uncomfortable around someone with HIV. | | |
| | People with HIV have only themselves to blame for getting HIV. | | | | People with HIV have only themselves to blame for getting HIV. | | ** |
| | People with HIV must have done something wrong to get it. | | | | People with HIV must have done something wrong to get it. | | |

| | |
|---|---|
| | People with HIV should feel ashamed about having HIV. |
| | I would be ashamed if someone in my family has HIV. |
| | If I was in public or private transport, I would not like to sit next to someone with HIV. |
| | I would not like to be friends with someone with HIV. |
| | I would not employ someone with HIV. |
| | I would not eat together with someone I knew had HIV. |
| | If a relative of mine became ill with HIV, I would not want to care for that person in my home. |

| | |
|---|---|
| People with HIV should feel ashamed about having HIV. | |
| I would be ashamed if someone in my family has HIV. | |
| If I was in public or private transport, I would not like to sit next to someone with HIV. | |
| I would not like to be friends with someone with HIV. | |
| I would not employ someone with HIV. | ✓ |
| I would not eat together with someone I knew had HIV. | ✓ |
| If a relative of mine became ill with HIV, I would not want to care for that person in my home. | |

| | |
|---|---|
| | I would not want to buy food from someone I know has HIV. |
| | If a teacher has HIV but is not sick, she should not be allowed to continue teaching in the school. |
| | I would not want someone with HIV to look after my child. |
| | I do not want to get too close to someone with HIV because I am afraid I might get infected with HIV. |
| | I would not want my child to play with a child who has HIV or whose parents have HIV. |
| N/A | **NEW ITEM** |
| | **NEW ITEM** |

| | |
|---|---|
| I would not want to buy food from someone I know has HIV. | |
| If a teacher has HIV, she should not be allowed to continue teaching in the school. | |
| I would not want someone with HIV to look after my child. | ✓ |
| I do not want to get too close to someone with HIV because I am afraid I might get infected with HIV. | ✓ |
| I would not want my child to play with a child who has HIV or whose parents have HIV. | ✓ |
| Women with HIV should not get pregnant | ✓ |
| Women with HIV should not be mothers. | |

| | | | | | | | |
|---|---|---|---|---|---|---|---|
| CASS[18] | Once you've had cancer you're never "normal" again. | | | | Once you have HIV, you're never normal "again" | | + |
| | Getting cancer means having to mentally prepare oneself for death. | | | | Getting HIV means having to mentally prepare oneself for death | ✓ | + |
| | Cancer devastates the lives of those it touches. | | | | HIV devastates the lives of those it touches | | + |
| ***Anticipated*** | | | | | | | |
| AHAS[20] | If people knew my HIV status, I would be treated badly by health workers. | If people knew my HIV status, I would be treated badly by health workers. | | | If I had HIV, I would be treated badly by health workers. | | |
| | If people knew my HIV status, I would lose my job, customers, or livelihood. | If people knew my HIV status, I would lose my job, customers, or livelihood. | | | If I had HIV, I would lose my job, customers, or livelihood. | ✓ | |
| | If people knew my HIV status, my family would not take care of me when I am sick. | If people knew my HIV status, my family would not take care of me when I am sick. | | | If I had HIV, my family would not take care of me when I am sick. | | |

| | | | | | | |
|---|---|---|---|---|---|---|
| | If people knew my HIV status, my family would reject or abandon me. | If people knew my HIV status, my family would reject or abandon me. | | | If I had HIV, my family would reject or abandon me. | ✓ |
| | If people knew my HIV status, I would be treated badly in my community. | If people knew my HIV status, I would be treated badly in my community. | | | If I had HIV, I would be treated badly in my community. | ✓ |
| | If people knew my HIV status, I would be physically abused by my partner. | If people knew my HIV status, I would be physically abused by my partner. | | | If I had HIV, I would be physically abused by my partner. | |
| | If people knew my HIV status, my relationship would end. | If people knew my HIV status, my relationship would end. + | ✓ | | If I had HIV, my relationship would end. | ✓ |
| | If people knew my HIV status, I would be shunned at social gatherings. | If people knew my HIV status, I would be shunned at social gatherings. | | | If I had HIV, I would be shunned at social gatherings. | ✓ |
| | If people knew my HIV status, I would lose friends. | If people knew my HIV status, I would lose friends. | ✓ | | If I had HIV, I would lose friends. | ✓ |

| | | | | | | | |
|---|---|---|---|---|---|---|---|
| | If people knew my HIV status, I would not be allowed to participate fully in the church or mosque. | If people knew my HIV status, I would not be allowed to participate fully in the church or mosque. | ✓ | | If I had HIV, I would not be allowed to participate fully in the church or mosque. | ✓ | |
| | If people knew my HIV status, I would lose respect/standing within my family or community. | If people knew my HIV status, I would lose respect/standing within my family or community. | | | If I had HIV, I would lose respect/standing within my family or community. | ✓ | |
| | If people knew my HIV status, people would gossip about me. | If people knew my HIV status, people would gossip about me. | | | If I had HIV, people would gossip about me. | | ** |
| ***Internalized*** | | | | | | | |
| | I have failed to live up to my own expectations by getting HIV. | I have failed to live up to my own expectations by getting HIV. | | | If I had HIV, I would feel that I have failed to live up to my own expectations by getting HIV. | ✓ | |
| HARSIH[19] | When I tell others I have HIV, I expect them to think less of me. | When I tell others I have HIV, I expect them to think less of me. | | | If I had HIV and told others that I had it, I would expect them to think less of me. | | |
| | I put myself down for becoming HIV+. | I put myself down for becoming HIV+. | ✓ | | If I had HIV, I would put myself down for becoming HIV+. | ✓ | |

| | | | | | | |
|---|---|---|---|---|---|---|
| | I put myself down for becoming HIV+. | Being HIV+ make me feel defective, like there is something wrong with me. | | | If I had HIV, it would make me feel defective, like there is something wrong with me. | ✓ |
| | I am ashamed that I'm HIV+. | I am ashamed that I'm HIV+. | ✓ | + | If I had HIV, I would be ashamed of my HIV+ status. | |
| | When others find out I am HIV+, I expect them to reject me. | When others find out I am HIV+, I expect them to reject me. | | | If I had HIV and others found out about it, I would expect them to reject me. | |
| | I struggle with feeling worthless because I have HIV. | I struggle with feeling worthless because I have HIV. | ✓ | | If I had HIV, I would struggle with feeling worthless because of my HIV status. | ✓ |
| | I am ashamed by my HIV symptoms. | | | | If I had HIV, I would hide my infection from others. | |
| | I hide my infection from others. | I hide my infection from others. | | | If I had HIV, I would hide my infection from others. | |
| | I have an overpowering dread that my HIV status will be revealed to others. | I am worried that my HIV status will be revealed to others. | ✓ | | If I had HIV, I would be worried that my HIV status will be revealed to others. | ✓ |
| | I accept myself as an HIV+ person. | I accept myself as an HIV+ person. | | | If I had HIV, I would accept myself as an HIV+ person. | |

| | | | | | | |
|---|---|---|---|---|---|---|
| | Having HIV makes me want to hide, disappear, or even die. | Having HIV make me want to hide, disappear, or even die. | | | If I had HIV, it would make me want to hide, disappear, or even die. | ✓ |
| HASI-P[17] | I felt that I did not deserve to live. | Having HIV makes me feel that I do not deserve to live. | ✓ | | If I had HIV, I would feel that I did not deserve to live. | |
| | I felt ashamed of having this disease. | I feel ashamed of having this disease. | ✓ | | If I had HIV, I would feel ashamed of having the disease. | ✓ |
| | I felt completely worthless. | Having HIV make me feel completely worthless. | ✓ | | If I had HIV, I would feel completely worthless. | ✓ |
| | I felt that I brought a lot of trouble to my family. | I feel that I brought a lot of trouble to my family. | | | If I had HIV, I would feel that I brought a lot of trouble to my family. | |
| | I felt that I am no longer a person. | I feel that I am no longer a member of my community. | ✓ | | If I had HIV, I would feel that I am no longer a member of my community. | ✓ |

** Suggested for Elimination During Cognitive Interview (Dhluo)
+ Added after Cognitive Interview
✓ Item kept in final item pool

| **Supplementary 6:** Summary of the HPV Stigmatizing Items with Sources and Modifications | | | | |
|---|---|---|---|---|
| **Item Origin** | **Item Adapted** | **Item Modification** | **Item Kept** | **Cognitive Interview** |
| | | ***Attitudes*** | | |
| HSS[15] | Getting HIV is a punishment for bad behavior. | Getting HPV is a punishment for bad behavior. | | D |
| | I would think less of someone if I found out the person has HIV. | I would think less of someone if I found out the person has HPV. | | |
| | I would be upset if someone with HIV moved in next door to me. | I would be troubled if someone with HPV moved in next door to me. | | |
| | I feel uncomfortable around someone with HIV. | I feel uncomfortable around someone with HPV. | | D |
| | People with HIV have only themselves to blame for getting HIV. | People with HPV have only themselves to blame for getting HPV. | ✓ | |
| | People with HIV must have done something wrong to get it. | People with HPV must have done something wrong to get it. | ✓ | |
| | People with HIV should feel ashamed about having HIV. | People with HPV should feel ashamed about having HPV. | | |
| | I would be ashamed if someone in my family has HIV. | I would be ashamed if someone in my family has HPV. | | |
| | I would not like to be friends with someone with HIV. | I would not like to be friends with someone with HPV. | | |
| | I would not eat together with someone I knew had HIV. | I would not eat together with someone I knew had HPV. | | |

| | | | | |
|---|---|---|---|---|
| | I would not eat together with someone I knew had HIV. | I would not want to buy food from someone I know has HPV. | | |
| | If a teacher has HIV but is not sick, she should not be allowed to continue teaching in the school. | If a teacher has HPV, she should not be allowed to continue teaching in the school. | | |
| | I would not want someone with HIV to look after my child. | I would not want someone with HPV to look after my child. | ✓ | D |
| | I do not want to get too close to someone with HIV because I am afraid I might get infected with HIV. | I do not want to get too close to someone with HPV because I am afraid I might get infected with HPV. | | D |
| | **NEW ITEM** | Women with HPV should not get pregnant. | ✓ | D |
| | **NEW ITEM** | Women with HPV should not be mothers. | | |
| CASS[18] | Once you've had cancer, you're never "normal" again. | Once you have HPV, you're never "normal" again. | ✓ | + |
| | Getting cancer means having to mentally prepare oneself for death. | Getting HPV means having to mentally prepare oneself for death. | | + |
| | Cancer devastates the lives of those it touches. | HPV devastates the lives of those it touches. | | + |
| ***Anticipated*** | | | | |
| HPV[21] | People would avoid me | If I had HPV, people would avoid me. | ✓ | |
| | People would think badly of me | If I had HPV, people would think badly of me. | | D |
| | People would blame me | If I had HPV, people would blame me. | ✓ | D |

| | | | | |
|---|---|---|---|---|
| | People would think I was unclean | If I had HPV, people would think I was unclean. | | E |
| | People would not want to be friends with me | f I had HPV, people would not want to be friends with me. | ✓ | |
| | People would not want to have a sexual relationship with me | If I had HPV, people would not want to have a sexual relationship with me. | | E |
| | People would be uncomfortable around me | If I had HPV, people would be uncomfortable around me. | ✓ | |
| | People would be angry with me | If I had HPV, people would be angry with me. | ✓ | D |
| | People would pity me | If I had HPV, people would pity me. | | |
| AHAS[20] | If people knew my HIV status, I would be treated badly by health workers. | If I had HPV, I would be treated badly by health workers. | ✓ | |
| | If people knew my HIV status, I would lose my job, customers, or livelihood. | If I had HPV, I would lose my job, customers, or livelihood. | ✓ | |
| | If people knew my HIV status, my family would reject or abandon me. | If I had HPV, my family would reject or abandon me. | | D |
| | If people knew my HIV status, I would be treated badly in my community. | If I had HPV, I would be treated badly in my community. | ✓ | |
| | If people knew my HIV status, I would be physically abused by my partner. | If I had HPV, I would be physically abused by my partner. | ✓ | |
| | If people knew my HIV status, my relationship would end. | If I had HPV, my relationship would end. | ✓ | |
| | If people knew my HIV status, I would lose friends. | If I had HPV, I would lose friends. | ✓ | |

| | | | | |
|---|---|---|---|---|
| | If people knew my HIV status, I would not be allowed to participate fully in the church or mosque. | If I had HPV, I would not be allowed to participate fully in the church or mosque. | ✓ | |
| | If people knew my HIV status, I would lose respect/standing within my family or community. | If I had HPV, I would lose respect/standing within my family or community. | ✓ | |
| | If people knew my HIV status, people would gossip about me. | If I had HPV, people would gossip about me. | | D |
| ***Anticipated Internalized*** | | | | |
| HARSIH[19] | Being HIV positive makes me feel defective, like there is something wrong with me. | Having HPV would make me feel like there is something wrong with me. | | |
| | I am ashamed that I'm HIV positive. | I would be ashamed to have HPV. | ✓ | |
| | I struggle with feeling worthless because I have HIV. | If I had HPV, I would struggle with feeling worthless because I have HPV. | ✓ | |
| | I hide my infection from others. | If I had HPV, I would hide my infection from others. | | |
| | I have an overpowering dread that my HIV status will be revealed to others. | If I had HPV, I would be very worried that my HPV status will be revealed to others. | | |
| | Having HIV makes me want to hide, disappear, or even die. | Having HPV would make me want to hide, disappear, or even die. | | |
| HASI-P[17] | I felt that I did not deserve to live. | If I had HPV, I would feel that I did not deserve to live. | ✓ | |

| | | | | |
|---|---|---|---|---|
| | I felt that I brought a lot of trouble to my family. | If I had HPV, I would feel that I brought a lot of trouble to my family. | ✓ | |
| | I felt that I am no longer a person. | If I had HPV, I would feel that I am no longer a woman. | ✓ | |
| HPV[21] | How responsible would you feel? | If I had HPV, I would feel responsible for getting HPV. | ✓ | D |
| | How ashamed would you feel? | If I had HPV, I would feel ashamed for getting HPV | ✓ | |
| | How embarrassed would you feel? | If I had HPV, I would feel embarrassed about having HPV | ✓ | |
| | How guilty would you feel? | If I had HPV, I would feel guilty for getting infected with HPV | ✓ | |
| | How disappointed in yourself would you feel? | If I had HPV, I would feel disappointed in myself for getting infected with HPV. | ✓ | |
| | How anxious would you feel? | I would feel anxious about living with HPV | | |
| | How scared would you feel? | I would be scared living with HPV. | | |
| | How angry would you feel? | I would feel angry about learning I had HPV | | |

E Suggested for Elimination During English Cognitive Interview

D Suggested for Elimination During Dhluo Cognitive Interview

+ Added after Cognitive Interview

✓ Item kept in final item pool

| **Supplementary 7:** Summary of the Cervical Cancer Stigmatizing Items with Sources and Modifications | | | | |
|---|---|---|---|---|
| **Item Origin** | **Item Adapted** | **Item Modification** | **Item Kept** | **Cognitive Interview** |
| ***Attitudes*** | | | | |
| CASS[18] | I would feel comfortable around someone with cancer | I would feel comfortable around someone with cervical cancer | | |
| | I would find it hard to talk to someone with cancer | I would find it hard to talk to someone with cervical cancer | ✓ | |
| | I would feel embarrassed discussing cancer with someone who had it | I would feel embarrassed discussing cervical cancer with someone who had it | ✓ | |
| | Once you've had cancer you're never "normal" again | Once you've had cervical cancer you're never "normal" again | | |
| | Getting cancer means having to mentally prepare oneself for death | Getting cervical cancer means having to mentally prepare oneself for death. | | ** |
| | Cancer usually ruins close personal relationships | Cervical cancer usually ruins close personal relationships | | |
| | Cancer devastates the lives of those it touches | Cervical cancer devastates the lives of those it touches | | |
| | I would feel irritated by someone with cancer | I would feel irritated by someone with cervical cancer. | X | ** |
| | I would feel angered by someone with cancer | I would feel angered by someone with cervical cancer | ✓ | |
| | I would try to avoid a person with cancer | I would try to avoid a person with cervical cancer | | |
| | A person with cancer is to blame for their condition | A person with cervical cancer is to blame for their condition | | |
| ***Anticipated Internalized*** | | | | |
| | **NEW ITEM** | If I had cervical cancer, I would be ashamed by my symptoms (bleeding, weight loss) | ✓ | |

| | | | | |
|---|---|---|---|---|
| HARSIH[19] | I am ashamed that I'm HIV+. | I would be ashamed to be diagnosed with cervical cancer. | ✓ | |
| | I put myself down for becoming HIV+. | I would blame myself for getting diagnosed with cervical cancer. | | |
| | Having HIV makes me want to hide, disappear, or even die. | Having cervical cancer would make me want to hide, disappear, or even die. | | ** |
| HASI-P[17] | I felt that I did not deserve to live. | If I had cervical cancer, I would feel that I did not deserve to live. | | ** |
| | I felt that I brought a lot of trouble to my family. | If I had cervical cancer, I would feel that I brought a lot of trouble to my family. | ✓ | |
| | I felt that I am no longer a person. | If I had cervical cancer, I would feel that I am no longer a woman. | ✓ | |
| HPV[21] | How embarrassed would you feel? | I would feel embarrassed about having cervical cancer | ✓ | |
| | How guilty would you feel? | I would feel guilty for having cervical cancer. | | ** |
| | How anxious would you feel? | (X) I would feel anxious about getting cervical cancer | | |
| | How scared would you feel? | I would be scared to have cervical cancer | | |
| | How angry would you feel? | I would feel angry if I learned I had cervical cancer | ✓ | |
| ***Anticipated*** | | | | |
| AHAS[20] | If people knew my HIV status, I would be treated badly by health workers. | If I had cervical cancer, I would be treated badly by health workers. | | |
| | If people knew my HIV status, I would lose my job, customers, or livelihood. | If I had cervical cancer, I would lose my job, customers, or livelihood. | | |
| | If people knew my HIV status, my family would reject or abandon me. | If I had cervical cancer, my family would reject or abandon me. | | |

| | | | |
|---|---|---|---|
| | If people knew my HIV status, I would be treated badly in my community. | If I had cervical cancer, I would be treated badly in my community. | |
| | If people knew my HIV status, I would be physically abused by my partner. | If I had cervical cancer, I would be physically abused by my partner. | |
| | If people knew my HIV status, my relationship would end. | If I had cervical cancer, my relationship would end. | ✓ |
| | If people knew my HIV status, I would be shunned at social gatherings. | If I had cervical cancer, I would be shunned at social gatherings. | ✓ |
| | If people knew my HIV status, I would lose friends. | If I had cervical cancer, I would lose friends. | ✓ |
| | If people knew my HIV status, I would not be allowed to participate fully in the church or mosque. | If I had cervical cancer, I would not be allowed to participate fully in the church or mosque. | |
| | If people knew my HIV status, I would lose respect/standing within my family or community. | If I had cervical cancer, I would lose respect/standing within my family or community. | ✓ |
| | If people knew my HIV status, people would gossip about me. | If I had cervical cancer, people would gossip about me. | ✓ |

** Suggested for Elimination During Cognitive Interview (Dhluo)

X Removed right after Cognitive Interview

✓ Item kept in final item pool

**Supplementary 8**: Confirmatory factor analysis and scale characteristics

| Model | Average (SD) Item Score | Average (95% CI) **Standardized** Loading |
|---|---|---|
| HPV | 0.77 (0.09) | 0.73 (0.68, 0.77) |
| CC | 0.9 (0.19) | 0.79 (0.69, 0.89) |
| HIVNEG | 0.69 (0.09) | 0.77 (0.7, 0.83) |
| HIVPOS | 0.63 (0.12) | 0.89 (0.67, 1.1) |

**Supplementary 9:** Fit statistics

| Model | CFI | RMSEA (90% CI) | SRMR | Weak invariance test |
|---|---|---|---|---|
| HPV Overall | 0.99 | 0.07 (0.06, 0.07) | 0.06 | |
| HPV Grouped | 0.99 | 0.08 (0.08, 0.09) | 0.08 | 0.01 |
| CC Overall | 0.99 | 0.09 (0.08, 0.1) | 0.07 | |
| CC Grouped | 0.98 | 0.12 (0.11, 0.13) | 0.09 | 0.12 |
| HIV- Overall | 0.99 | 0.11 (0.11, 0.12) | 0.08 | |
| HIV- Grouped | 0.98 | 0.14 (0.13, 0.14) | 0.1 | <0.01 |
| HIV+ Overall | 0.98 | 0.12 (0.09, 0.14) | 0.09 | |
| HIV+ Grouped | 0.98 | 0.14 (0.11, 0.16) | 0.12 | 0.14 |
